# Normative ranges for auditory brainstem response wave I amplitude: A potential diagnostic indicator of cochlear deafferentation

**DOI:** 10.1101/2025.11.13.25340158

**Authors:** Sean D Kampel, Garnett P McMillan, Anne E Heassler, Nicole K Whittle, Haley A Szabo, Naomi F Bramhall

## Abstract

**Purpose:** While cochlear synaptopathy has limited impact on auditory thresholds, there is increasing evidence that cochlear deafferentation is associated with auditory perceptual deficits. However, there is currently no means for diagnosing synaptopathy or deafferentation in individual patients. The objectives of this study were to establish normative ranges for auditory brainstem response (ABR) wave I amplitude, a measure sensitive to synaptopathy in animals, in a population at low risk for synaptopathy and then compare a population at high-risk for synaptopathy to the normative ranges.

**Methods:** The low-risk sample consisted of 169 non-Veteran young adults with normal audiograms, minimal self-reported noise exposure history, and no auditory complaints (tinnitus, decreased sound tolerance, or speech perception in noise difficulty). ABR wave I amplitude normative ranges were generated for 2, 4, and 8 kHz tonebursts and were statistically adjusted for sex and average distortion product otoacoustic emission (DPOAE) levels. Ninety-one military Veterans with normal audiograms and at least one auditory complaint were included in the high-risk comparison sample.

**Results:** While the DPOAE-adjusted ABR normative ranges were effective at distinguishing between the low– and high-risk samples, the results also indicated that adjusting the ABR normative ranges for OHC dysfunction may not be necessary and could be problematic. ABR normative ranges that were adjusted only for sex were able to differentiate between the low– and high-risk samples, with 51% of the high-risk sample falling below the normative ranges for a 105 dB peSPL 8 kHz toneburst.

**Conclusions:** In patients with normal audiograms, sex-specific ABR wave I amplitude normative ranges can be used by clinicians to identify patients with high degrees of cochlear deafferentation.

## Introduction

One longstanding challenge in clinical audiology is how to address the paradox of patients reporting auditory complaints despite having normal hearing thresholds. These complaints include tinnitus, difficulty hearing in background noise or complex listening environments, and decreased sound tolerance (DST). In a review of 235,091 health records from military Veterans, Billings et al. (2018) found that approximately 1 in 10 Veterans seeking hearing health care had normal hearing thresholds. They also noted that this is likely an underestimate because normal hearing test results were not consistently submitted to the data records repository. Similarly, an analysis of data collected from 2783 participants in the Beaver Dam Offspring Study found that 12% reported hearing difficulty despite having normal hearing thresholds in both ears (Tremblay et al., 2015). One potential etiology for auditory complaints in the context of a normal audiogram is cochlear synaptopathy, damage to the synapses between the inner hair cells (IHCs) and the afferent auditory nerve fibers.

Cochlear synaptopathy was first reported in a mouse model following a two-hour exposure to an octave band noise at 100 dB SPL that induced a temporary threshold shift (Kujawa & Liberman, 2009). While otoacoustic emissions and auditory brainstem response (ABR) thresholds fully recovered post-noise exposure, suprathreshold ABR wave 1 amplitudes remained reduced even at 8 weeks post-exposure. Histological analysis indicated that approximately 50% of cochlear synapses were lost in the cochlear frequency region above the noise band despite preservation of the IHCs and outer hair cells (OHCs). Since that seminal study, noise-induced cochlear synaptopathy has been demonstrated in several other animal models, including rat (Hickox et al., 2017), guinea pig (Lin et al., 2011), chinchilla (Hickox et al., 2017), and rhesus macaque (Valero et al., 2017). Age-related cochlear synaptopathy has also been observed in animal models (Parthasarathy & Kujawa, 2018; Schmiedt et al., 1996; Sergeyenko et al., 2013). Human temporal bone data suggests that age– and noise exposure-related synaptopathy also occur in humans (Wu et al., 2019; Wu et al., 2021). The predicted perceptual consequences of this synaptopathy include tinnitus, hyperacusis, and difficulty understanding speech in background noise (Kujawa & Liberman, 2015). These data suggest that synaptopathy-related auditory complaints are likely a common problem impacting audiology patients and identifying this problem in individual patients could help with the development of treatment options.

Unfortunately, there is currently no method for diagnosing synaptopathy in living humans because synaptopathy can only be confirmed through post-mortem temporal bone analysis. However, several auditory physiological measures appear to be sensitive to synaptopathy in animal models (Kujawa & Liberman, 2009; Shaheen et al., 2015; Valero et al., 2018). One of these measures, ABR wave I amplitude, assesses the combined output of the auditory nerve fibers (Hashimoto et al., 1981) and is a non-invasive metric that many audiology clinics are already equipped to obtain. As would be expected with cochlear synaptopathy, reductions in ABR wave I amplitude are associated with aging (Carcagno & Plack, 2020; Grant et al., 2020; Johannesen et al., 2019; Konrad-Martin et al., 2012) and military noise exposure (Bramhall et al., 2017; Bramhall et al., 2021), even after accounting for OHC function. Several studies have demonstrated that tinnitus is associated with reduced ABR wave I amplitudes (Bramhall et al., 2018; Bramhall et al., 2019; Bramhall et al., 2023; Gu et al., 2012; Schaette & McAlpine, 2011; Valderrama et al., 2018), while others have shown that reduced ABR wave I amplitude is associated with speech-in-noise perception difficulty (Bramhall, Buran, & McMillan, 2025; reviewed in Bramhall & McMillan, 2024). It is important to note that ABR wave I amplitude, as a measure of the signal at the level of the auditory nerve, cannot differentiate between cochlear synaptopathy, IHC loss, and spiral ganglion cell loss and for that reason we refer to it as an indicator of cochlear deafferentation (Bramhall, 2021). However, human temporal bone studies indicate that age-related synapse loss exceeds the loss of IHCs and spiral ganglion cells (Wu et al., 2019), noise exposure is more likely to damage cochlear synapses than IHCs (Wu et al., 2021), and synapses are lost following noise exposure prior to loss of spiral ganglion cells (Kujawa & Liberman, 2009). Taken together, these data suggest that cochlear deafferentation is driven primarily by synapse loss.

Although ABR wave I amplitude has potential as a diagnostic indicator of cochlear deafferentation, clinicians are currently unable to use ABR wave I amplitude for this purpose because it isn’t clear what constitutes a “normal” ABR wave I amplitude. Normative ranges are commonly used in audiological testing to identify patients with abnormal auditory function. A key aspect of normative ranges is that they are developed by sampling from a population expected to be at low risk of dysfunction. For example, pure-tone thresholds are referenced to normative threshold data that was collected from adults assumed to have normal hearing (Fletcher & Wegel, 1922; Gatlin & Dhar, 2021).

Use of ABR wave I amplitude as an indicator of cochlear deafferentation is also complicated by potential impacts of sex and OHC dysfunction on the measurement. Mitchell et al (1989) found that women had larger wave I amplitudes than men, potentially due to sex-related differences in head size. Computational modeling by Verhulst et al. (2018) showed a non-linear relationship between ABR wave I amplitude and OHC dysfunction as a function of ABR stimulus intensity level, where at lower intensity levels OHC dysfunction results in decreased wave I amplitudes while at higher intensity levels OHC dysfunction leads to increased wave I amplitudes. These findings suggest that it may be important to account for sex and OHC dysfunction when using ABR wave I amplitude for diagnostic purposes. In the past, human studies of synaptopathy typically addressed this issue by requiring normal auditory thresholds for study participation and/or by statistically adjusting ABR wave I amplitudes for either pure tone thresholds or distortion product otoacoustic emissions (DPOAEs; e.g., Bramhall et al., 2017; Prendergast et al., 2017; Skoe & Tufts, 2018).

The objectives of the current study were 1) to establish normative ranges for ABR wave I amplitude in a population at low risk for cochlear synaptopathy and 2) to compare these normative ranges to ABR wave I amplitude measurements collected from a population at high risk of synaptopathy to determine how well the normative ranges can distinguish between populations at low versus high risk of synaptopathy. To account for the potential impacts of sex and OHC dysfunction on ABR wave I amplitude, separate normative ranges were developed for males and females and the normative ranges were statistically adjusted for average DPOAE level.

## Methods

### Participants

Participants were recruited from the VA Portland Health Care System, local colleges and universities, and the surrounding Portland community. All participants provided informed consent prior to any study activities and were paid for their time. The research protocol and all study procedures were approved by the Institutional Review Board of the VA Portland Health Care System.

### Normative low-risk sample

169 young non-Veteran adults aged 18 to 35 years (49 males, 120 females) were included in the normative low-risk sample. This sample was expected to be at low-risk for cochlear synaptopathy due to their young age, minimal noise exposure history, and lack of auditory complaints (tinnitus, speech-in-noise perception difficulty, or DST). All participants had clinically normal hearing thresholds (≤ 20 dB from 0.25-8 kHz), no audiometric air-bone gaps greater than 15 dB and no more than one air-bone gap of 15 dB from 0.5-4 kHz, and normal tympanograms (±50 daPa, compliance 0.3-1.5 ml) in at least one ear. Individuals with a history of military service, any reported history of firearm use, or a history of concussion were not included in the sample.

All low-risk participants completed the occupational and recreational sections of the Lifetime Exposure to Noise and Solvents Questionnaire (LENS-Q; Griest-Hines et al., 2021). The LENS-Q was scored as described in Griest-Hines et al. Results from a previous study found a mean LENS-Q score of 4.1 (standard deviation 0.8) for a similar sample of young non-Veterans with normal hearing and minimal noise exposure history (Bramhall et al., 2021). Based on these findings, potential participants with LENS-Q scores ≥ 5 were judged to have too much noise exposure history to be included in the sample.

Low risk participants were also asked to complete a questionnaire that included several questions about auditory complaints. The question, “Have you experienced ringing, roaring, or buzzing in the ears or head (tinnitus) that lasts for at least 5 minutes?” was used to evaluate tinnitus perception. Potential low-risk participants who responded, “yes” to this question were excluded. Self-reported speech perception in noise difficulty was assessed by asking potential participants to rate their perceived difficulty hearing in several situations (taken from the Hearing section of the Tinnitus and Hearing Survey [THS; Henry et al., 2015]). These situations include noisy or crowded places, understanding what people are saying on TV or in movies, understanding people with soft voices, and understanding what is being said in group conversations. Individuals who reported hearing difficulty in two or more of these situations were excluded. Potential low-risk participants were also asked to respond to the following question from the Sound Tolerance section of the THS: “Over the last week, sounds were too loud or uncomfortable for me when they seemed normal to others around me.” Individuals who responded “yes” to this question were asked to provide two examples of sounds that were too loud or uncomfortable for them. If an individual’s responses were judged by the examiner (a licensed audiologist) to be consistent with DST, they were excluded from the sample.

Wideband middle ear muscle reflex (MEMR) normative ranges developed from data obtained from 142 of these participants and envelope following response (EFR) normative ranges based on data from 154 of these participants were previously reported (Bramhall, McMillan, et al., 2025; Heassler et al., 2025).

### High-risk comparison sample

Data was collected from a separate sample of 91 military Veterans aged 23 to 49 years (56 males, 35 females) who were expected to be at high risk for cochlear synaptopathy due to their history of military noise exposure and report of at least one auditory complaint (tinnitus, difficulty understanding speech-in-noise, and/or DST) for the purpose of comparing to the ABR normative ranges. All high-risk participants met the same audiometric and tympanometric inclusion criteria as the low-risk participants and did not have any history of concussion.

Potential high-risk participants were asked the same questions about auditory complaints as the participants from the low-risk sample. Only those that reported tinnitus, hearing difficulty in at least one of the situations from the THS, and/or DST were included. In addition, all high-risk participants completed the Speech, Spatial and Qualities of Hearing Scale (SSQ12; Noble et al., 2013), a 12-item questionnaire that asks about hearing difficulty in a variety of listening situations. Average scores range from 0 to 10, with lower scores indicating greater difficulty. High-risk participants who reported tinnitus were also asked to complete the Tinnitus Functional Index (TFI; Meikle et al., 2012), a 25-item questionnaire that asks about the social and emotional impacts of an individual’s tinnitus to assess how it impacts their quality of life. Total scores range from 0 to 100, with higher scores indicating more severity.

Wideband MEMR data from 83 of the high-risk participants and EFR data from 81 of the high-risk participants were reported previously (Bramhall, McMillan, et al., 2025; Heassler et al., 2025).

### Procedures

All testing was performed by a licensed audiologist.

### Distortion product otoacoustic emissions (DPOAEs)

To assess OHC function, DPOAEs were measured in the test ear using a custom system with an Etymotic Research ER-10X probe microphone system and EMAV software (Neely & Liu, 1993). A primary frequency sweep (DP-gram) was obtained in 1/3-octave increments from 1-8 kHz and 1/6-octave increments from 9-16 kHz with a frequency ratio of *f_2_/f_1_*=1.2 and *L_1_/L_2_* levels of 65/55 dB forward pressure level (FPL). Using in-the-ear calibration, the voltage was adjusted to set *L_1_* and *L_2_* to the desired levels. At each test frequency, the following measurement based stopping rules were used: 1) either 48 s of artifact free data were collected, 2) data were collected until the noise floor was below –30 dB FPL, or 3) data was collected until the signal-to-noise ratio was ≥ 30 dB.

For statistical adjustment of the normative ranges, DPOAE levels were averaged from either 3-8 kHz or 3-16 kHz. The rationale for measuring DPOAEs in the extended high frequencies (up to 16 kHz) was based on evidence that there are large contributions to ABR wave I amplitude from auditory nerve fibers at frequencies higher than the ABR stimulus frequency (Don & Eggermont, 1978). However, because most commercial DPOAE systems cannot reliably measure frequencies above 8 kHz, normative ranges adjusted for average DPOAE level from 3-8 kHz were also calculated. If the DPOAE level measured at a particular frequency was < –20 dB FPL, that frequency was not included in the DPOAE average.

The ER10X allows for FPL calibration, which increases the precision of the DPOAE measurement (Bharadwaj et al., 2019). Other than the differences in measurement error, dB values in FPL and sound pressure level (SPL) are equivalent. Most commercially available DPOAE systems provide measurements in dB SPL. Therefore, to avoid confusion, DPOAE level is reported in dB SPL in all figures and tables in this report.

### Auditory brainstem response (ABR)

ABR data were obtained using an Intelligent Hearing Systems Smart EP system (Miami, FL) and ER-3A ultra-shielded insert earphones. Stimuli consisted of 2, 4, and 8 kHz alternating polarity tonebursts with a Blackman envelope and a repetition rate of 11.1/sec presented at 95 dB peak equivalent (pe) SPL and 105 dB peSPL. Amplifier settings were configured with artifact rejection set to 31.0 µV, amplifier gain set to 100,000, the high-pass filter set to 10 Hz, and the low-pass filter set to 1500 Hz. Stimulus durations were 3000 µs for 2 kHz, 2000 µs for 4 kHz, and 1000 µs for 8 kHz tonebursts. An ear canal electrode (tiptrode) was placed in the test ear (non-inverting active electrode) and Ambu Neuroline 720 surface electrodes (Columbia, MD) were placed on the high forehead (Fz, inverting reference electrode) and low forehead (FpZ, ground). Weaver and Company Nuprep skin prep gel (Aurora, CO) was used to abrade the skin in preparation for electrode placement. All electrode impedances were below 5 kΩ except for one participant where impedances were ≤ 5.4 kΩ. For each stimulus frequency, 2048 sweeps were collected for recordings at 95 dB peSPL and 1024 sweeps were collected for recordings at 105 dB peSPL to minimize noise exposure. At least two waveforms were obtained for each stimulus condition. Additional waveforms were collected if the consistency of the first two waveforms was judged to be poor by the licensed audiologist performing the testing based on peak latencies, amplitudes, and morphology, following Hall (2007; pages 214-215). All collected ABR waveforms that were not excluded due to a clear testing problem such as participant movement, electrical artifact, or a problem with the transducer were included in the final analysis.

ABR testing was conducted in a sound booth where the lights and power were turned off during testing to avoid power line interference. Research participants were situated in a comfortable recliner, asked to close their eyes, and encouraged to sleep during the testing if possible. If participants were unable to sleep, they were asked to remain still and quiet during testing. ABR testing typically took one hour to complete, including setup and instructions. Equipment calibrations were performed twice per year using a Brüel & Kjær (Darmstadt, Germany) head and torso simulator (model 4128) with artificial ear simulators (4158-C, 4159-C), and a 3160-A-042 digitizer. **Figure 1** shows how the intensity level was calculated for a 105 dB peSPL 4 kHz toneburst.

**Figure 1.**
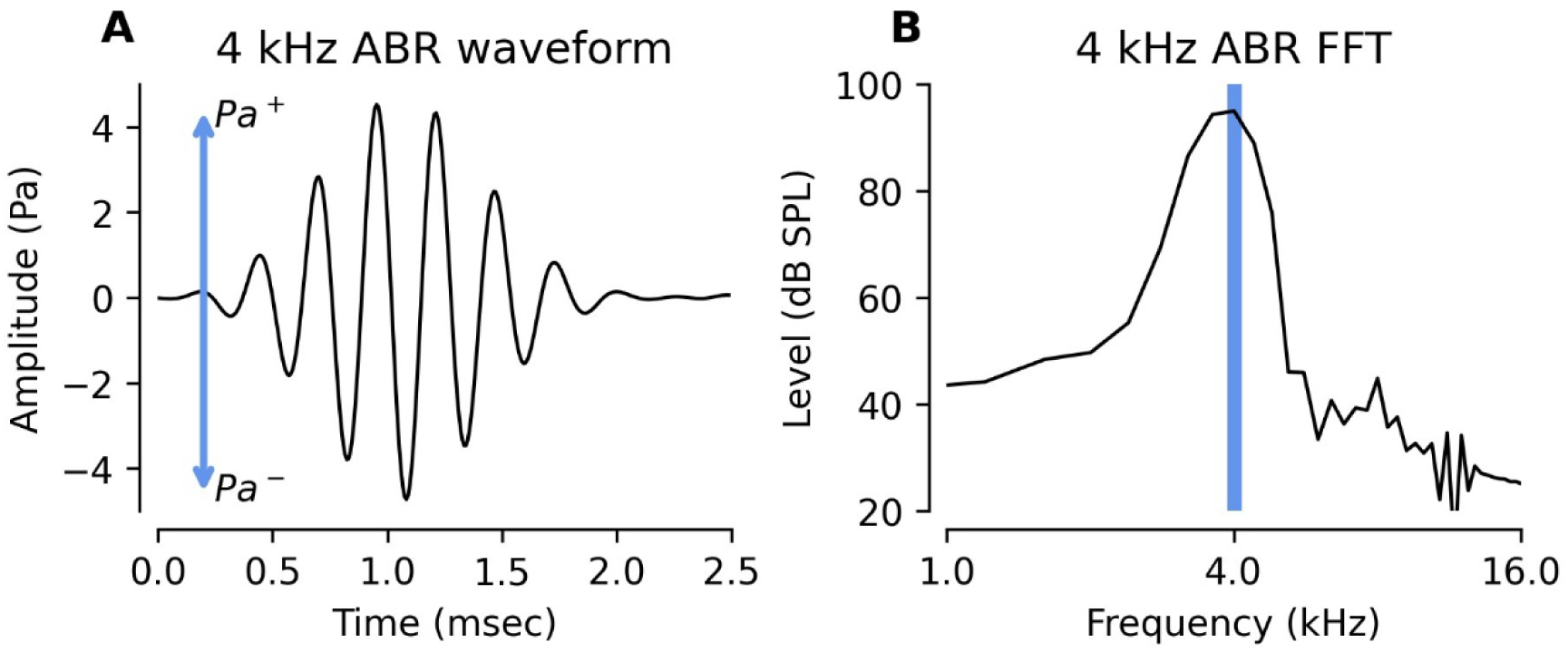
Example recorded ABR stimulus and fast Fourier transform. A 105 dB peak equivalent SPL (peSPL) 4 kHz ABR toneburst was presented through an ER-3A transducer to a head and torso simulator with an artificial ear simulator. **(A)** Recorded stimulus waveform. Blue arrow represents the difference between the positive and negative peaks that were used to calculate the dB peSPL. **(B)** Frequency spectrum of the ABR stimulus. Blue bar indicates the stimulus frequency (4 kHz).

Wave I amplitude was calculated as the difference in voltage between the wave I peak and the following trough (see **Figure 2**). ABR waveforms were initially analyzed by custom peak-picking software (Buran, 2015) and then evaluated by two licensed audiologists, who made corrections to the automated peak picking as necessary. Any disagreements between the two audiologists were resolved by a third licensed audiologist.

**Figure 2.**
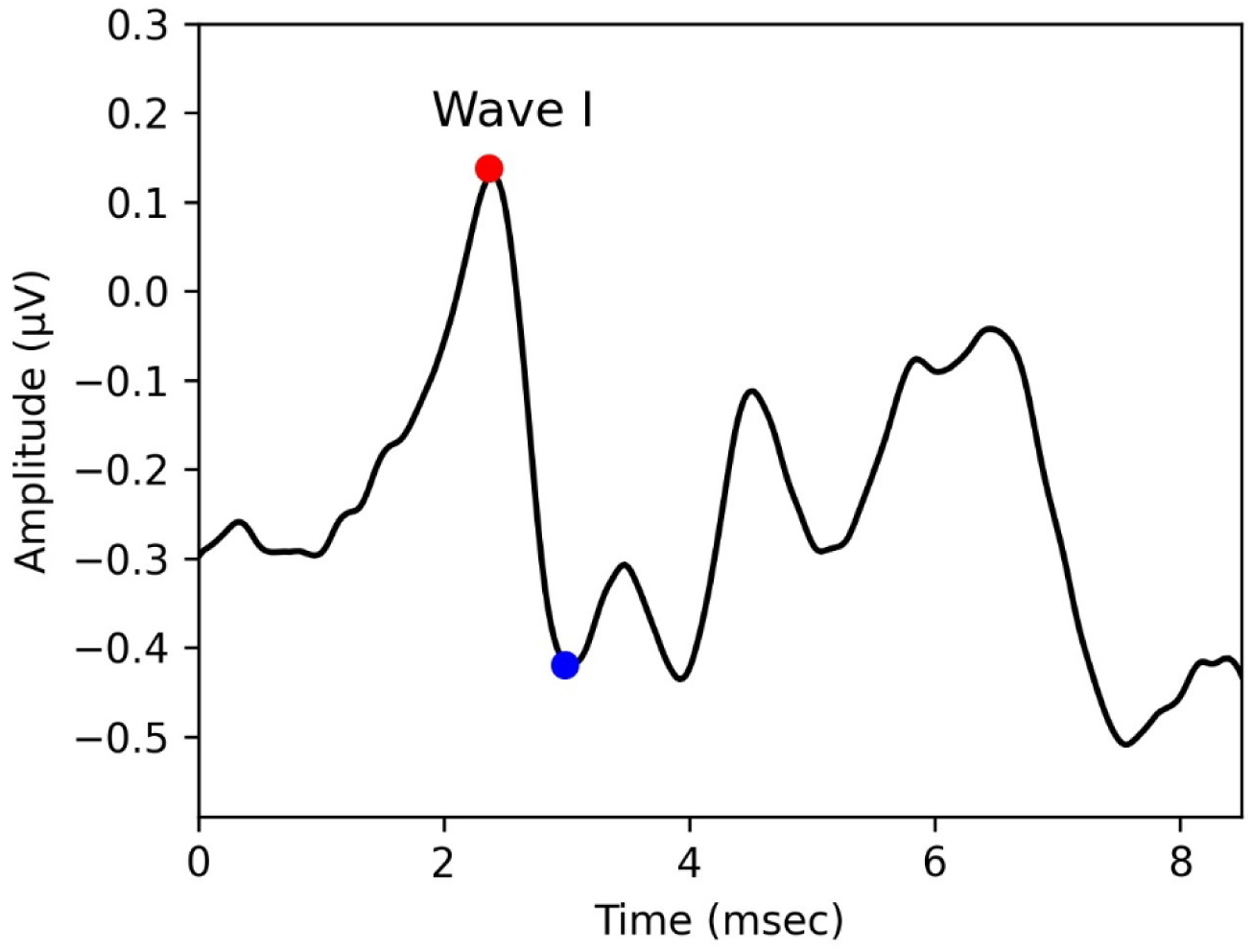
Example ABR response waveform. Plot shows how ABR wave I amplitude was calculated as the difference between the wave I peak (red dot) and the following trough (blue dot).

### Generation of normative ranges

Normative ranges provide specific percentiles of a reference population’s measurement distribution that can be used to identify patients who have unusual measurements relative to a ‘healthy normal’ population (Wright & Royston, 1999). Normative ranges are determined from measurements collected from a sample of healthy individuals. Conditional normative ranges specify population percentiles that vary based on a patient characteristic, such as age. For example, age-specific normative ranges are often generated by collecting measurements from a large number of healthy individuals across different age categories, and then estimating the normative ranges separately for each age category. An alternative approach for generating conditional normative ranges requires statistical modeling of the relationship between the population percentiles and the characteristic of interest. This approach is called quantile regression and involves fitting regression models to estimate the 10^th^ and 90^th^ percentiles in a sample of healthy individuals with varying levels of the conditional variable. While standard regression models focus on the mean of the measurement distribution, quantile regression is more flexible because it allows for modeling of any percentiles of the data distribution based on the covariates. It is recommended that at least 125 participants are used for estimating this type of non-parametric normative ranges (Linnet, 1987).

In this analysis, only data from the low-risk sample was used to generate the normative ranges. For each ABR stimulus condition, the 10^th^ and 90^th^ percentiles of the ABR wave I amplitude distribution were conditioned on participant sex and measured DPOAE levels averaged from either 3-16 kHz or 3-8 kHz. Non-linear relationships between the percentiles and DPOAE levels were evaluated, but they performed poorly in the sample and did not provide clear advantages over a linear model. The final model estimated population percentiles based on average DPOAE level, participant sex, and the interaction between average DPOAE level and sex. Because the purpose of developing these normative ranges was to identify individuals likely to have high degrees of cochlear deafferentation, particular attention should be given to the lower bounds (10^th^ percentile) of the normative ranges because measurements falling below the lower bound suggest an abnormally small ABR wave I amplitude, consistent with cochlear deafferentation.

## Results

ABR wave I amplitude normative ranges for males and females, adjusted for average DPOAE level from 3-16 kHz or 3-8 kHz, are shown in **Figures 3-6** (blue shaded region). The 10^th^ percentile of the low-risk sample is indicated by the lower bounds of the normative ranges. By definition, if ABR data was collected from a new sample drawn from the same population at low risk for synaptopathy, 10% of those individuals would be expected to fall below the 10^th^ percentile of the normative ranges. Because cochlear deafferentation is expected to reduce ABR wave I amplitude, the lower bound of the ABR wave I amplitude normative range is what will be used to identify individuals with high degrees of deafferentation. Having an ABR wave I amplitude that falls below the lower bound indicates an abnormally small wave I amplitude, consistent with a significant amount of deafferentation. The ABR wave I amplitudes at the upper and lower bounds of the normative ranges are shown in **Table 1** and **Table 2** for DPOAEs averaged from 3-16 kHz and 3-8 kHz, respectively.

**Figure 3.**
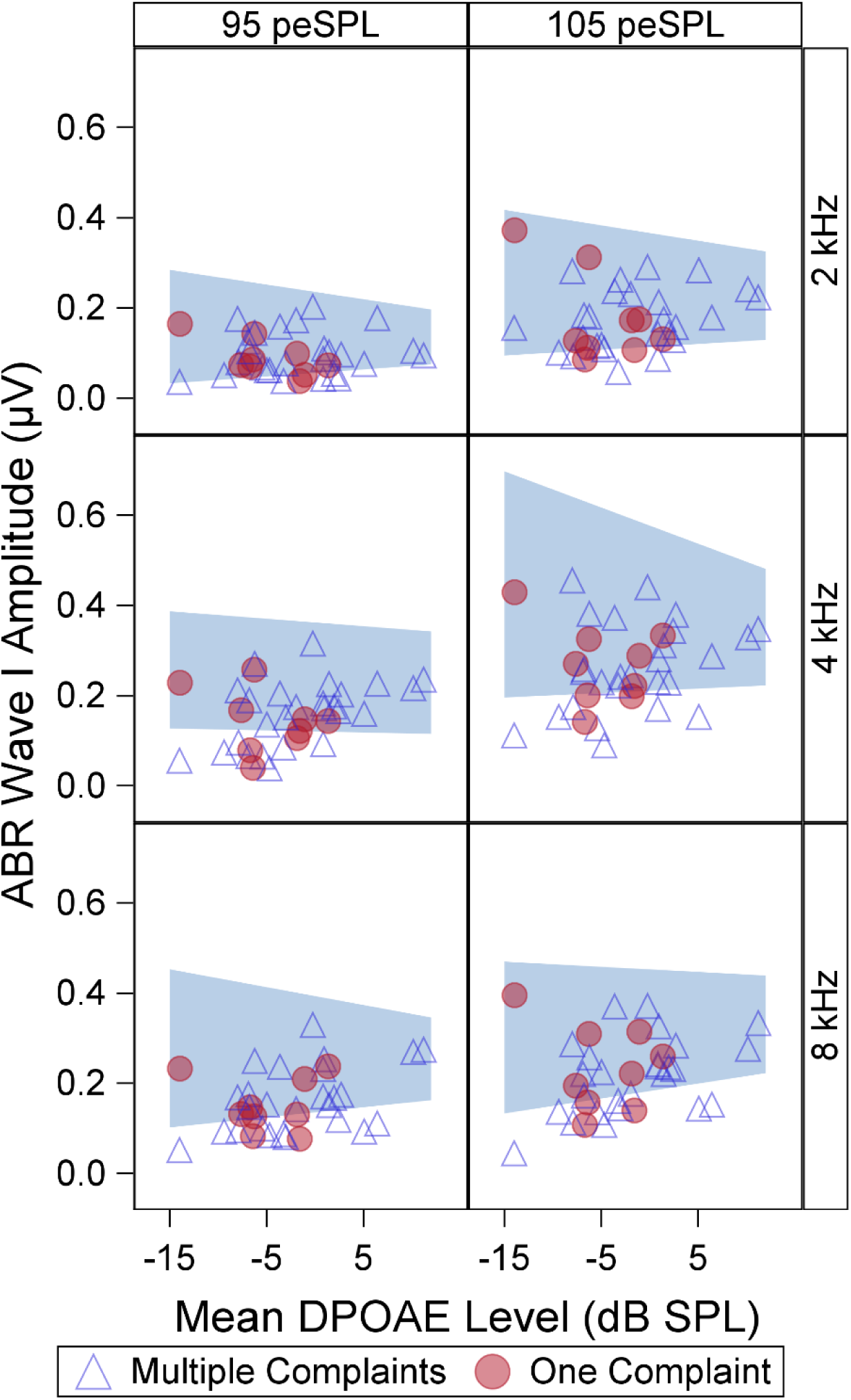
Female ABR wave I amplitude normative ranges adjusted for average DPOAE level from 3-16 kHz. Each panel shows the normative range for a different ABR stimulus with 95 dB peSPL stimuli in the left column and 105 dB peSPL stimuli in the right column. Blue shaded region indicates the DPOAE-adjusted normative range. Symbols show how ABR data from high-risk participants compares to the normative ranges (open blue triangles for individuals with one auditory complaint, filled red circles for individuals with multiple auditory complaints).

**Figure 4.**
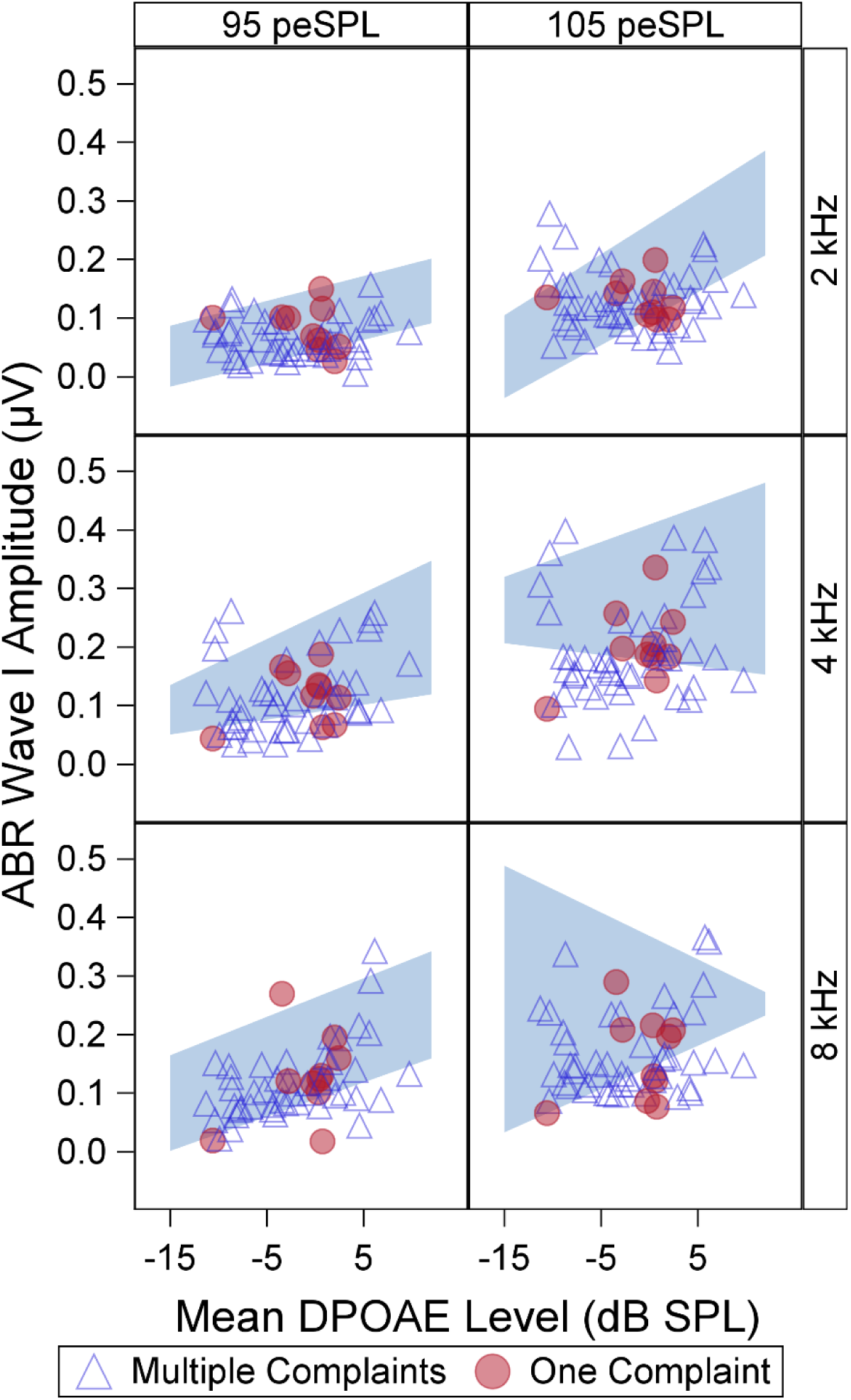
Male ABR wave I amplitude normative ranges adjusted for average DPOAE level from 3-16 kHz. Each panel shows the normative range for a different ABR stimulus with 95 dB peSPL stimuli in the left column and 105 dB peSPL stimuli in the right column. Blue shaded region indicates the DPOAE-adjusted normative range. Symbols show how ABR data from high-risk participants compares to the normative ranges (filled red circles for individuals with one auditory complaint, open blue triangles for individuals with multiple auditory complaints).

**Figure 5.**
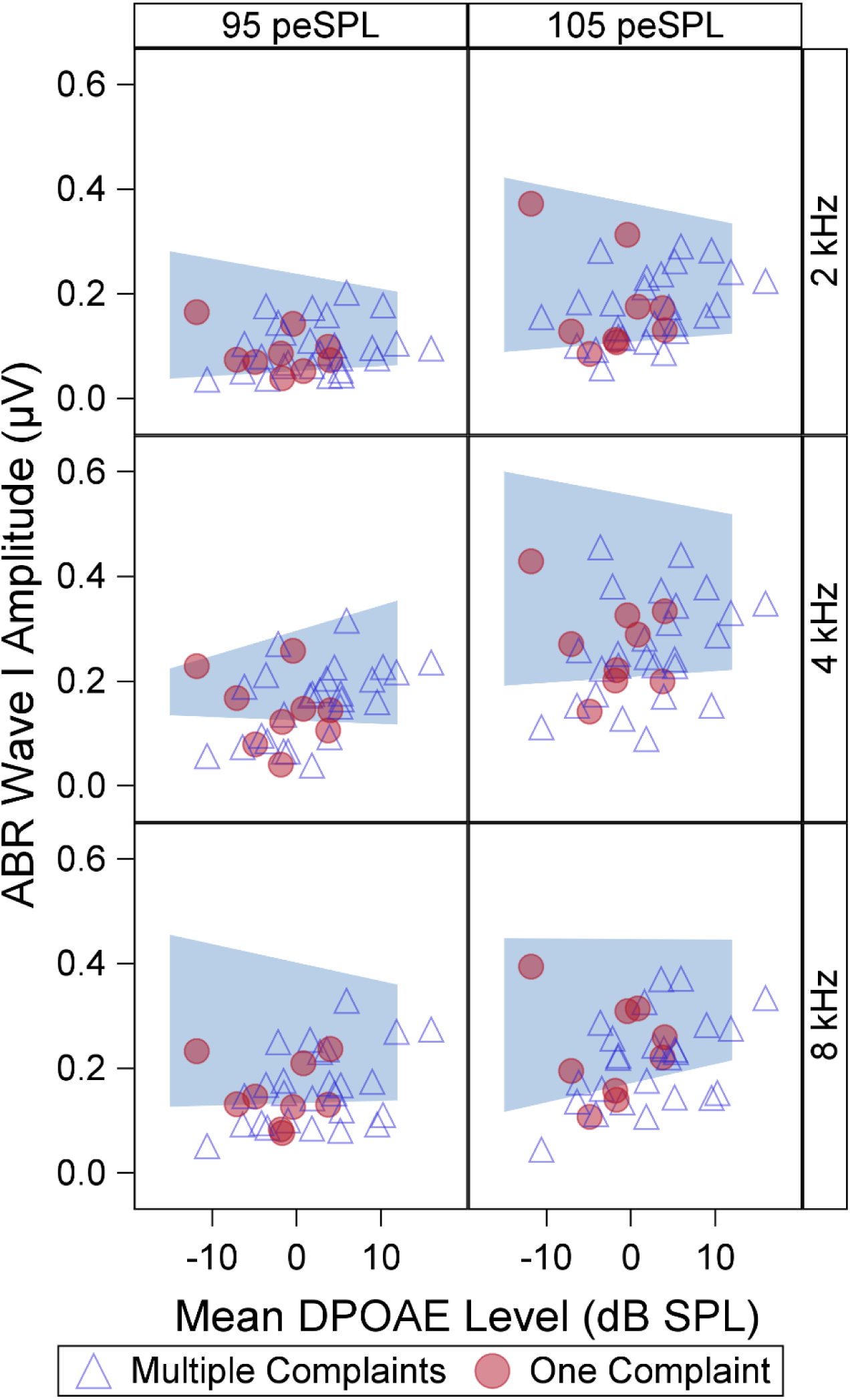
Female ABR wave I amplitude normative ranges adjusted for average DPOAE level from 3-8 kHz. Each panel shows the normative range for a different ABR stimulus with 95 dB peSPL stimuli in the left column and 105 dB peSPL stimuli in the right column. Blue shaded region indicates the DPOAE-adjusted normative range. Symbols show how ABR data from high-risk participants compares to the normative ranges (filled red circles for individuals with one auditory complaint, open blue triangles for individuals with multiple auditory complaints).

**Figure 6.**
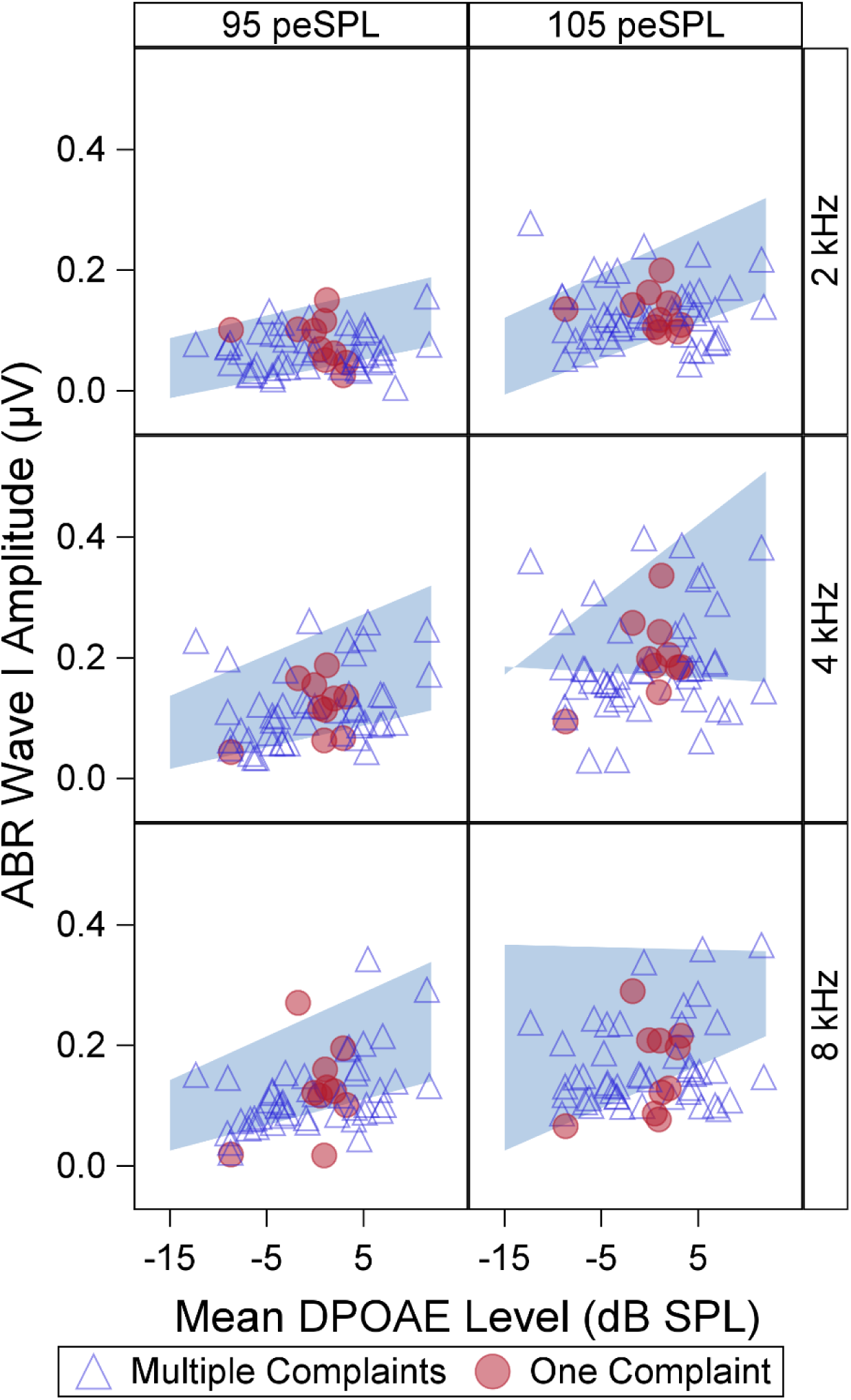
Male ABR wave I amplitude normative ranges adjusted for average DPOAE level from 3-8 kHz. Each panel shows the normative range for a different ABR stimulus with 95 dB peSPL stimuli in the left column and 105 dB peSPL stimuli in the right column. Blue shaded region indicates the DPOAE-adjusted normative range. Symbols show how ABR data from high-risk participants compares to the normative ranges (filled red circles for individuals with one auditory complaint, open blue triangles for individuals with multiple auditory complaints).

**Table 1.**
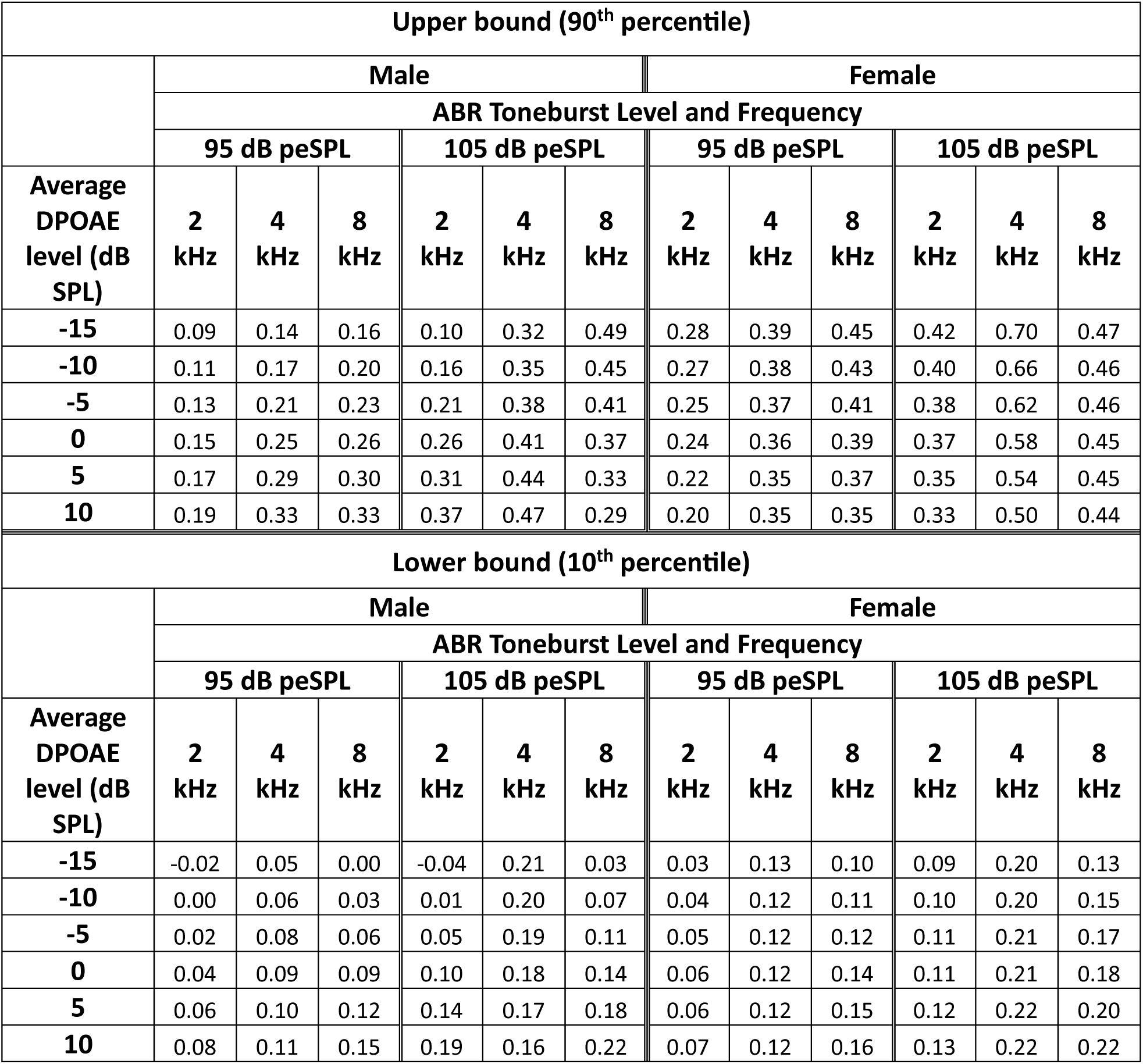
Lower (10^th^ percentile) and upper bounds (90^th^ percentile) for ABR wave I amplitude normative ranges adjusted for sex and average DPOAE levels from 3-16 kHz.

| Upper bound (90<sup>th</sup> percentile) |  |  |  |  |  |  |  |  |  |  |  |  |
| --- | --- | --- | --- | --- | --- | --- | --- | --- | --- | --- | --- | --- |
|  | Male |  |  |  |  |  | Female |  |  |  |  |  |
|  | ABR Toneburst Level and Frequency |  |  |  |  |  |  |  |  |  |  |  |
|  | 95 dB peSPL |  |  | 105 dB peSPL |  |  | 95 dB peSPL |  |  | 105 dB peSPL |  |  |
| Average DPOAE level (dB SPL) | 2 kHz | 4 kHz | 8 kHz | 2 kHz | 4 kHz | 8 kHz | 2 kHz | 4 kHz | 8 kHz | 2 kHz | 4 kHz | 8 kHz |
| -15 | 0.09 | 0.14 | 0.16 | 0.10 | 0.32 | 0.49 | 0.28 | 0.39 | 0.45 | 0.42 | 0.70 | 0.47 |
| -10 | 0.11 | 0.17 | 0.20 | 0.16 | 0.35 | 0.45 | 0.27 | 0.38 | 0.43 | 0.40 | 0.66 | 0.46 |
| -5 | 0.13 | 0.21 | 0.23 | 0.21 | 0.38 | 0.41 | 0.25 | 0.37 | 0.41 | 0.38 | 0.62 | 0.46 |
| 0 | 0.15 | 0.25 | 0.26 | 0.26 | 0.41 | 0.37 | 0.24 | 0.36 | 0.39 | 0.37 | 0.58 | 0.45 |
| 5 | 0.17 | 0.29 | 0.30 | 0.31 | 0.44 | 0.33 | 0.22 | 0.35 | 0.37 | 0.35 | 0.54 | 0.45 |
| 10 | 0.19 | 0.33 | 0.33 | 0.37 | 0.47 | 0.29 | 0.20 | 0.35 | 0.35 | 0.33 | 0.50 | 0.44 |
| Lower bound (10<sup>th</sup> percentile) |  |  |  |  |  |  |  |  |  |  |  |  |
|  | Male |  |  |  |  |  | Female |  |  |  |  |  |
|  | ABR Toneburst Level and Frequency |  |  |  |  |  |  |  |  |  |  |  |
|  | 95 dB peSPL |  |  | 105 dB peSPL |  |  | 95 dB peSPL |  |  | 105 dB peSPL |  |  |
| Average DPOAE level (dB SPL) | 2 kHz | 4 kHz | 8 kHz | 2 kHz | 4 kHz | 8 kHz | 2 kHz | 4 kHz | 8 kHz | 2 kHz | 4 kHz | 8 kHz |
| -15 | -0.02 | 0.05 | 0.00 | -0.04 | 0.21 | 0.03 | 0.03 | 0.13 | 0.10 | 0.09 | 0.20 | 0.13 |
| -10 | 0.00 | 0.06 | 0.03 | 0.01 | 0.20 | 0.07 | 0.04 | 0.12 | 0.11 | 0.10 | 0.20 | 0.15 |
| -5 | 0.02 | 0.08 | 0.06 | 0.05 | 0.19 | 0.11 | 0.05 | 0.12 | 0.12 | 0.11 | 0.21 | 0.17 |
| 0 | 0.04 | 0.09 | 0.09 | 0.10 | 0.18 | 0.14 | 0.06 | 0.12 | 0.14 | 0.11 | 0.21 | 0.18 |
| 5 | 0.06 | 0.10 | 0.12 | 0.14 | 0.17 | 0.18 | 0.06 | 0.12 | 0.15 | 0.12 | 0.22 | 0.20 |
| 10 | 0.08 | 0.11 | 0.15 | 0.19 | 0.16 | 0.22 | 0.07 | 0.12 | 0.16 | 0.13 | 0.22 | 0.22 |

**Table 2.**
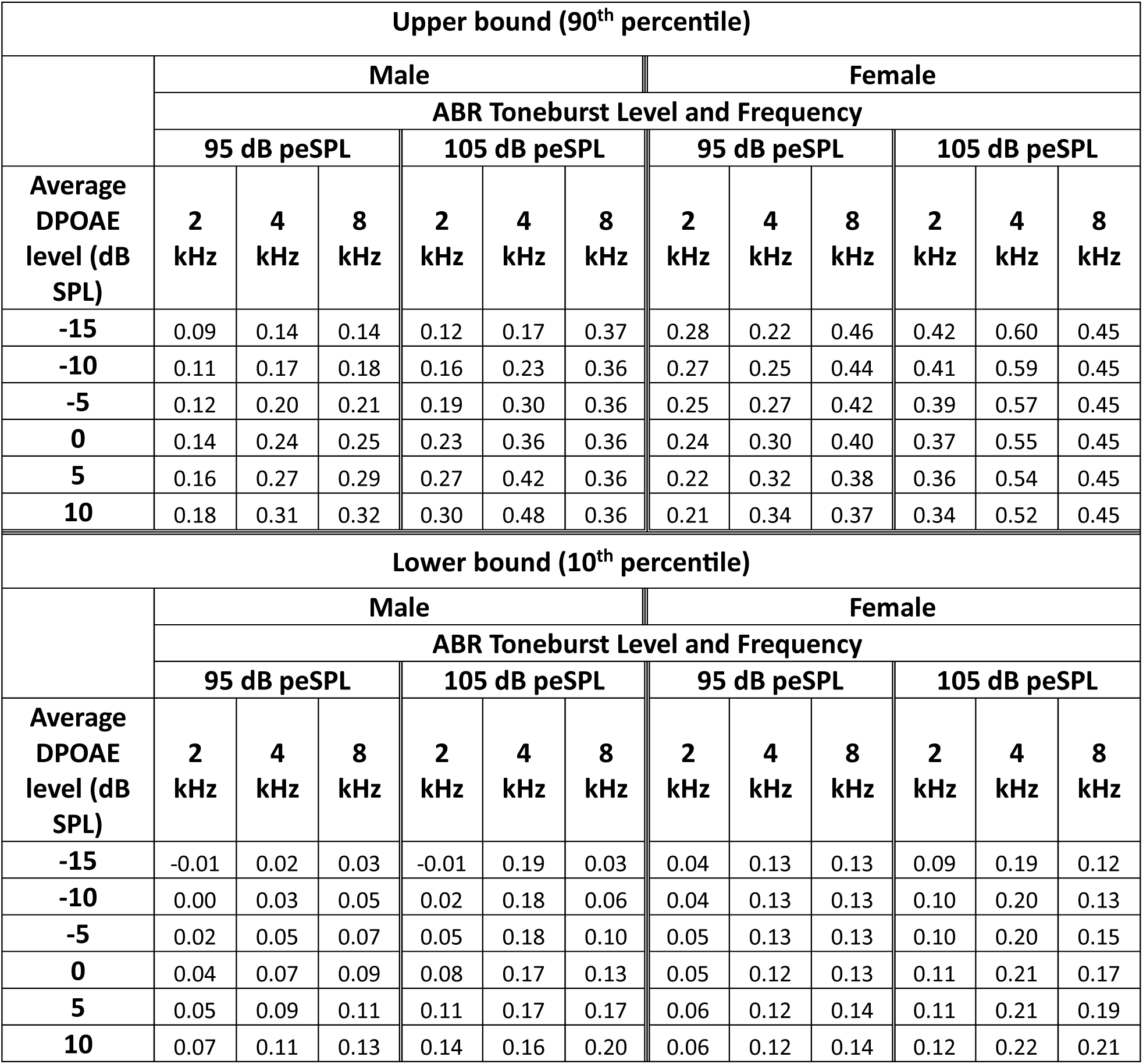
Lower (10^th^ percentile) and upper bounds (90^th^ percentile) for ABR wave I amplitude normative ranges adjusted for sex and average DPOAE levels from 3-8 kHz.

### Stimulus frequency and level effects on normative ranges

Normative ranges for each ABR stimulus condition, adjusted for average DPOAE level from 3-16 kHz, are shown for females and males in **Figure 3** and **Figure 4**, respectively. Across frequencies, increasing the ABR stimulus level from 95 to 105 dB peSPL generally raised both the lower (10^th^ percentile) and the upper bounds (90^th^ percentile) of the normative range and broadened the normative range, increasing the range of wave I amplitudes that would be considered normal. As ABR stimulus frequency increased from 2 kHz to 4 kHz, the lower bounds of the normative range increased (by 0.03 to 0.10 µV). At 95 dB peSPL, the lower bounds of the normative ranges increased as frequency increased from 4 kHz to 8 kHz for males and females with robust DPOAEs (average DPOAE level ≥ 5 dB SPL), but to a smaller degree (by 0.02-0.04 µV). At 105 dB peSPL, the lower bounds of the normative range only increased from 4 kHz to 8 kHz for males with robust DPOAEs.

### Sex effects on normative ranges

The lower and upper bounds of the normative ranges (adjusted for average DPOAE level from 3-16 kHz) tended to be higher for females than for males (by up to 0.38 µV). Sex differences were greatest for the 105 dB peSPL 4 kHz stimulus for poorer DPOAEs. However, there were some exceptions to this trend of an upward shift in the normative ranges for females relative to males (e.g., for males with robust DPOAEs for a 105 dB peSPL 2 kHz stimulus).

### Effects of DPOAE adjustment on normative ranges

In the female normative ranges adjusted for average DPOAE level from 3-16 kHz, the lower bounds were relatively flat (i.e., they did not change much with average DPOAE level), although there was a slight upward shift as DPOAEs became more robust for all stimuli except for the 95 dB peSPL 4 kHz stimulus (**Figure 3**). In contrast, the male normative ranges adjusted for average DPOAE level from 3-16 kHz (**Figure 4**) generally showed a clear upward slope of the lower bound as DPOAEs became more robust. The only exception was the male normative range for a 105 dB peSPL 4 kHz stimulus, where the slope of the lower bound was reversed and increased slightly as DPOAEs become poorer.

Similar trends were observed for the normative ranges adjusted for average DPOAE level from 3-8 kHz (**Figure 5** and **Figure 6**). The lower bounds for the normative ranges tended to be slightly lower when adjusting for average DPOAE level from 3-8 kHz versus average DPOAE level from 3-16 kHz (**Table 1** compared to **Table 2**).

### Auditory complaints reported in high-risk sample

**Table 3** summarizes the auditory complaints reported by the high-risk participants. Most participants reported more than one auditory complaint, with tinnitus and speech perception difficulty being the most frequently co-occurring complaints. Many participants reported all three auditory complaints (n=22, 24%). Participants with multiple complaints who reported tinnitus had higher mean TFI scores, indicating greater tinnitus severity, than participants who reported tinnitus alone. Mean SSQ-12 scores were poorer for participants who reported speech perception difficulty than for those who did not.

**Table 3.** Report of auditory complaints in the high-risk sample.

|  | # of males | # of females | Age | TFI score | SSQ score |
| --- | --- | --- | --- | --- | --- |
| Tinnitus only | 3 | 1 | 42 (5.8) | 20.1 (4.8) | 8.4 (0.6) |
| Speech perception only | 5 | 8 | 38.0 (5.6) |  | 6.7 (1.3) |
| DST only | 1 | 0 | 23 |  | 7.6 |
| Tinnitus + speech perception | 32 | 14 | 38.2 (6.1) | 42.3 (15.6) | 6.0 (1.4) |
| Speech perception + DST | 3 | 2 | 39.6 (3.5) |  | 6.1 (1.5) |
| Tinnitus + speech perception + DST | 12 | 10 | 37.5 (7.0) | 45.0 (11.8) | 5.8 (1.5) |

The last 3 columns show mean values with the standard deviation in parentheses. DST – decreased sound tolerance; TFI – Tinnitus Functional Index; SSQ – Speech, Spatial, and Qualities of Hearing scale

### Comparison of high-risk sample to ABR normative ranges

ABR data from individual high-risk participants is overlayed on the ABR normative ranges in **Figures 3-6** (symbols). Individuals whose ABR wave I amplitude fell below the normative ranges would be considered abnormal and likely to have a high degree of cochlear deafferentation. **Table 4** lists the percentage of high-risk participants who fell below the normative range for each stimulus condition and DPOAE average. The normative ranges for a 105 dB peSPL 4 kHz toneburst performed the best at differentiating between the low and high-risk populations, with 43% of the high-risk sample falling below the lower bound of the normative range when adjusting for DPOAE average from 3-16 kHz and 36% falling below the lower bound when using the DPOAE average from 3-8 kHz. The normative ranges for a 105 dB peSPL 8 kHz toneburst also performed well, with 35% of high-risk participants falling below the normative range when adjusting for average DPOAE level from 3-16 kHz and 33% falling below the normative range when adjusting for average DPOAE level from 3-8 kHz. The normative ranges for a 95 dB peSPL 2 kHz toneburst performed the poorest, with only 18% of high-risk participants falling below the normative range, regardless of the DPOAE average that was used. For all stimulus conditions and DPOAE averages, few of the high-risk participants (2-4%) exceeded the upper bounds of the normative range. There were no clear differences in the distributions of ABR wave I amplitudes between participants with one auditory complaint and those with multiple complaints.

**Table 4.**
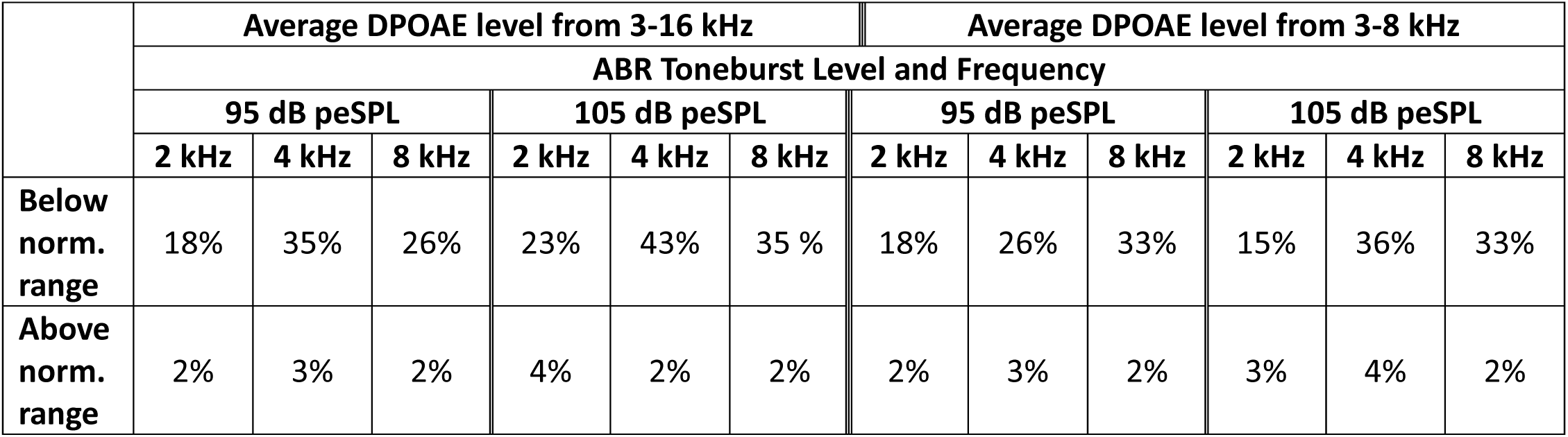
Percentage of high-risk sample falling above or below ABR wave I amplitude normative ranges adjusted for sex and average DPOAE level.

|  | Average DPOAE level from 3-16 kHz |  |  |  |  |  | Average DPOAE level from 3-8 kHz |  |  |  |  |  |
| --- | --- | --- | --- | --- | --- | --- | --- | --- | --- | --- | --- | --- |
|  | ABR Toneburst Level and Frequency |  |  |  |  |  |  |  |  |  |  |  |
|  | 95 dB peSPL |  |  | 105 dB peSPL |  |  | 95 dB peSPL |  |  | 105 dB peSPL |  |  |
|  | 2 kHz | 4 kHz | 8 kHz | 2 kHz | 4 kHz | 8 kHz | 2 kHz | 4 kHz | 8 kHz | 2 kHz | 4 kHz | 8 kHz |
| Below norm. range | 18% | 35% | 26% | 23% | 43% | 35 % | 18% | 26% | 33% | 15% | 36% | 33% |
| Above norm. range | 2% | 3% | 2% | 4% | 2% | 2% | 2% | 3% | 2% | 3% | 4% | 2% |

## Discussion

### DPOAE-adjusted normative ranges for a 105 dB peSPL 4 kHz stimulus performed best at differentiating between the low– and high-risk samples

Consistent with our expectations, many individuals from the high-risk sample had ABR wave I amplitudes that fell below the lower bounds of the normative ranges, suggesting significant cochlear deafferentation. More high-risk individuals fell below the lower bound of the DPOAE-adjusted normative ranges for a 105 dB peSPL 4 kHz stimulus (36-43%) than for any other stimulus, suggesting that this stimulus may be particularly good at differentiating between populations at low versus high risk of cochlear deafferentation. This is consistent with the results of Bramhall et al. (2017), which showed greater mean differences in ABR wave I amplitude between Veterans reporting high levels of noise exposure and non-Veteran controls for a 110 dB peSPL 4 kHz toneburst than for 1, 3, or 6 kHz tonebursts or for lower intensity 4 kHz tonebursts (80, 90, or 100 dB peSPL). Given that the high-risk sample were Veterans with a history of military noise exposure, they may be most likely to have deafferentation in the 4 kHz cochlear region because Veterans are more likely to have an audiometric notch at 4 kHz than at any other frequency (Wilson & McArdle, 2013). Higher ABR stimulus levels may perform better at differentiating low versus high-risk populations because at higher stimulus levels the distribution of wave I amplitudes widens and the normative range becomes broader, enhancing differences between normal and reduced responses.

### ABR normative ranges differed for males and females

Normative ranges for females were shifted upwards compared to normative ranges for males. This is consistent with the findings of Mitchell et al. (1989) that ABR wave I amplitudes tend to be larger for females than for males, potentially due to sex-related differences in head size. This suggests that it is important to have separate normative ranges for males and females, otherwise males would be more likely to fall below the normative ranges than females as a function of their sex rather than their degree of deafferentation.

### DPOAE adjustment may not be necessary

The relatively flat lower bounds observed in the female ABR normative ranges suggest that, in individuals with clinically normal hearing, subclinical OHC dysfunction may have little impact on suprathreshold ABR wave I amplitudes. This suggests that it may not be necessary to statistically adjust the normative ranges for average DPOAE level. This is consistent with recent findings in mice with varying degrees of synaptopathy and outer hair cell dysfunction, where the ability to predict synapse numbers from ABR wave 1 amplitude measurements was not improved by adjusting for DPOAEs (Buran et al., 2025). This calls into question why the same pattern of flat lower bounds was not observed in the male ABR normative ranges. There is no reason to expect that the impact of subclinical OHC dysfunction on ABR wave I amplitude would be different for males than for females. One possibility is that there was a stronger correlation between average DPOAE level and ABR wave I amplitude among male low-risk participants than female low-risk participants, with lower DPOAE levels being associated with lower ABR wave I amplitudes. This association could result from greater degrees of noise exposure-related cochlear damage in the male versus the female low-risk participants. Mean LENS-Q scores were almost identical for male and female low-risk participants (3.36 for females and 3.40 for males), suggesting similar degrees of noise exposure. However, data from mice indicates that males are more susceptible to noise-induced cochlear damage due to lower estrogen levels (Shuster et al., 2021), which could lead to greater cochlear damage in males than females even when noise exposure histories are similar. This may also explain why the slope of the lower bound of the male normative ranges was not consistent across all the stimuli but instead reversed for the 105 dB peSPL 4 kHz stimulus. Computational modeling suggests that the impacts of OHC dysfunction on ABR wave I amplitude vary with stimulus level (Verhulst et al., 2018) and this non-linear relationship may have unpredictable impacts on the DPOAE adjustment of the normative ranges.

Overall, it appears that DPOAE adjustment of the ABR normative ranges may not be necessary or desirable. Consequently, sex-adjusted ABR wave I amplitude normative ranges for 2, 4, and 8 kHz were recalculated without the DPOAE adjustment (**Figure 7**). The sex-adjusted normative ranges were calculated with the same quantile regression approach as the DPOAE adjusted normative ranges, but without the DPOAE predictor. When the high-risk sample was compared with these normative ranges, 37% of participants fell below the normative range for the 105 dB peSPL 4 kHz stimulus. The 105 dB peSPL 8 kHz normative range, unadjusted for DPOAEs, performed best at differentiating between the two populations, with 51% of high-risk participants falling below the lower bound. Overall, this suggests that high frequency suprathreshold ABR wave I amplitudes can distinguish between populations at low versus high risk for synaptopathy. The lower and upper bounds for the sex-adjusted ABR normative ranges are shown in **Table 5** and the percentage of high-risk participants falling outside of these normative ranges are shown in **Table 6**.

**Figure 7.**
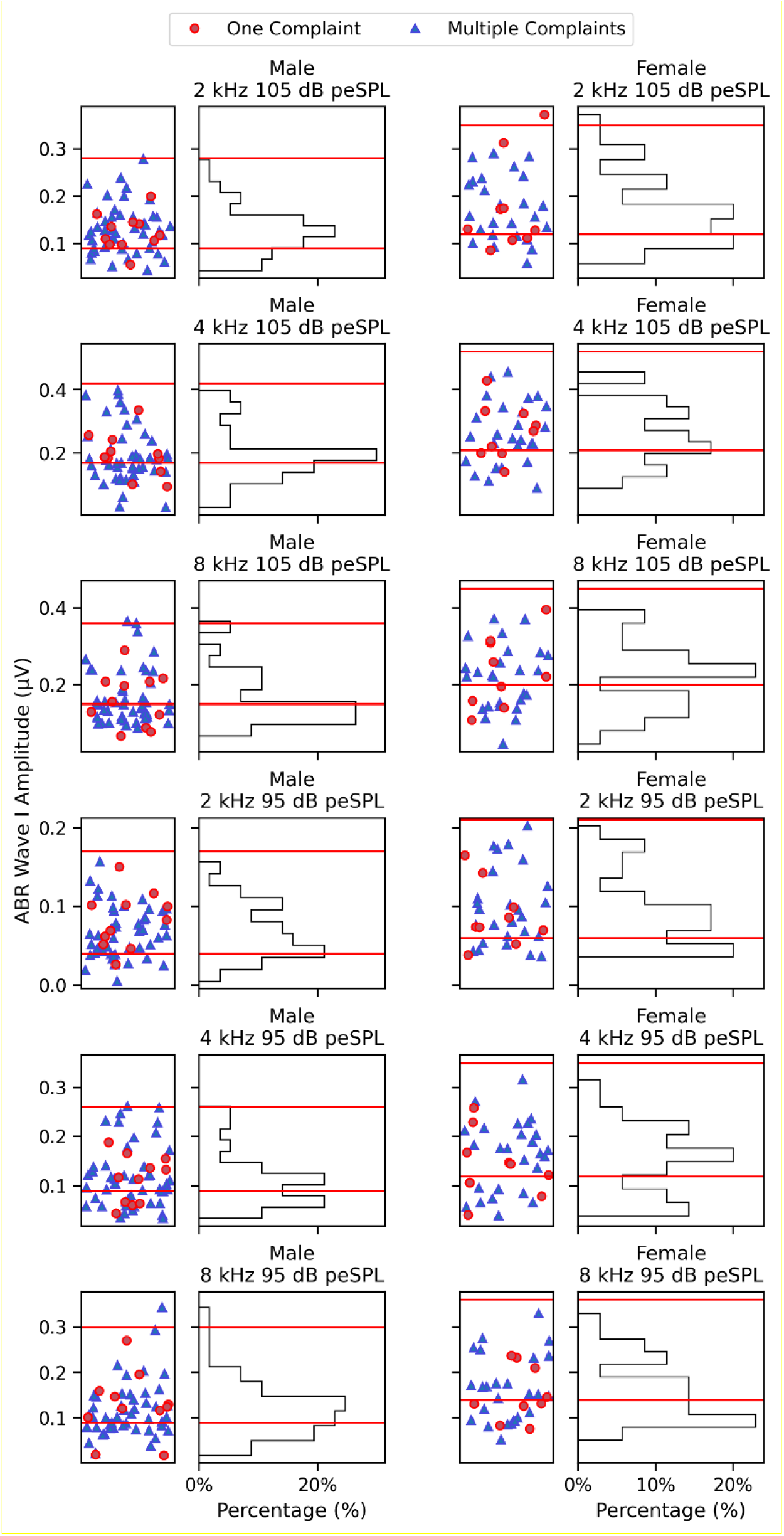
ABR wave I amplitude normative ranges adjusted for sex. Each pair of panels shows the normative range for a different ABR stimulus for males on the left-hand side and females on the right-hand side. In the scatterplots, the ABR wave I amplitude for each high-risk participant is represented as a symbol (red circle for one auditory complaint, blue triangle for multiple complaints). Histograms show the distribution of ABR wave I amplitudes for all high-risk participants. The upper (90^th^ percentile) and lower (10^th^ percentile) bounds of the normative ranges are indicated by red horizontal lines on both types of plots.

**Table 5.**
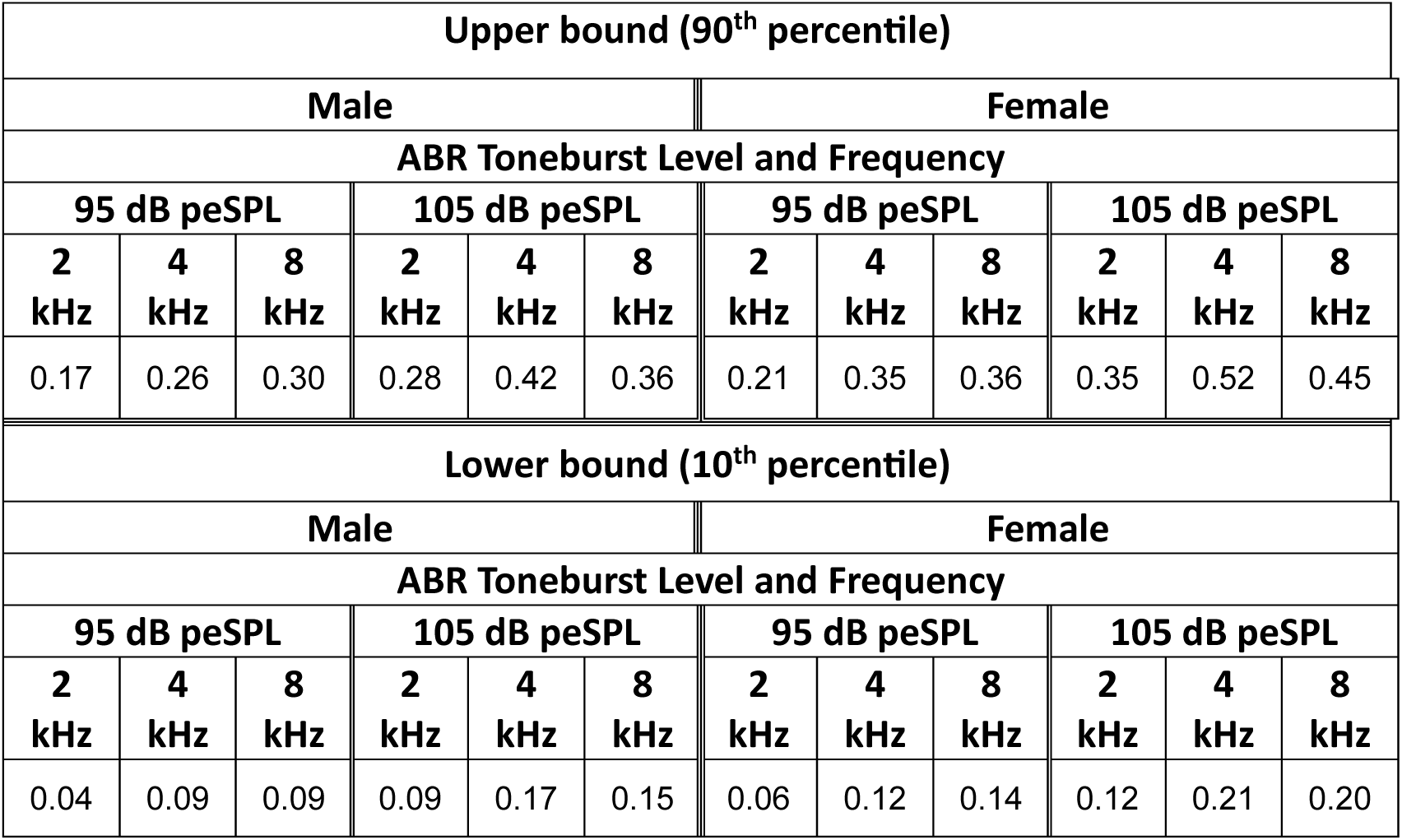
Lower (10^th^ percentile) and upper bounds (90^th^ percentile) for ABR wave I amplitude normative ranges adjusted for sex.

**Table 6.** Percentage of high-risk sample falling above or below ABR wave I amplitude normative ranges adjusted for sex.

|  | ABR Toneburst Level and Frequency |  |  |  |  |  |
| --- | --- | --- | --- | --- | --- | --- |
|  | 95 dB peSPL |  |  | 105 dB peSPL |  |  |
|  | 2 kHz | 4 kHz | 8 kHz | 2 kHz | 4 kHz | 8 kHz |
| **Below normative range** | 22% | 32% | 38% | 24% | 37% | 51% |
| **Above normative range** | 0% | 0% | 1% | 1% | 0% | 1% |

### Future clinical use of ABR normative ranges

The 105 dB peSPL 8 kHz ABR normative ranges for males and females shown in **Figure 7** can be used by clinicians in the future when evaluating patients who have auditory complaints despite normal audiometric thresholds. Patients who fall below the lower bound of the normative range are likely to have significant cochlear deafferentation. A diagnosis of cochlear deafferentation will allow the clinician to counsel the patient about the source of their auditory complaints and will guide the selection of treatment options. Most audiology clinics are already equipped to perform ABR testing, which should simplify clinical adoption of ABR wave I amplitude testing for this purpose. Prior to initiating ABR wave I amplitude testing, it is recommended that each clinic collect ABR data from a low-risk sample using their own ABR equipment. The characteristics of this validation sample should parallel those for the low-risk sample used in this study in terms of age, reported noise exposure, and lack of auditory complaints. If the equipment is appropriately calibrated, approximately 10% of a new low-risk sample should fall below the lower bound of the normative ranges and approximately 10% should surpass the upper bound.

Two other physiological indicators of cochlear synaptopathy, the wideband MEMR and the EFR, have been identified in animal models (Shaheen et al., 2015; Valero et al., 2018) and can be measured in humans. We previously demonstrated that wideband MEMR normative ranges developed in a low-risk sample and tested in a high-risk sample (participants overlapping with those described in this report) are unable to distinguish between populations at low versus high risk of synaptopathy (Bramhall, McMillan, et al., 2025). Although similarly developed EFR normative ranges were able to differentiate between the low– and high-risk samples (Heassler et al., 2025), recent data suggests that ABR wave I amplitude may perform better than the EFR at identifying individuals with deafferentation. In mice with age-related and noise-induced synaptopathy, ABR wave 1 amplitude was better at predicting synapse numbers than the EFR stimulus (modulated at 110 Hz) typically used in human synaptopathy studies (Buran et al., 2025). In addition, it is more common for audiology clinics to already have the equipment and expertise necessary for collecting ABR measurements than for collecting EFR measurements and limitations of the EFR measurement that could impact its use as a diagnostic indicator of deafferentation, such as the possibility of confounding from post-auricular muscle artifact and central gain (discussed in Heassler et al., 2025), should not impact ABR wave I amplitude.

## Conclusions

ABR wave I amplitude normative ranges were generated in a population at low risk for cochlear synaptopathy (young adults with normal hearing thresholds, minimal noise exposure history, and no auditory complaints). In females, the lower bounds of the normative ranges were relatively unimpacted by average DPOAE level, suggesting that it may not be necessary to measure and adjust for DPOAEs in patients with normal hearing thresholds. However, ABR normative ranges were impacted by sex, with smaller ABR wave I amplitudes required for males to fall below the normative ranges than for females. This indicates that it is important to use sex-specific normative ranges for ABR wave I amplitude. Sex-specific normative ranges, unadjusted for DPOAEs, for a 105 dB peSPL 8 kHz ABR toneburst were the most effective at differentiating the low-risk normative population from a population at high risk for synaptopathy (military Veterans with normal hearing thresholds and auditory complaints), with 51% of the high-risk sample falling below the normative ranges. This suggests that the ABR normative ranges are effective at identifying individuals with high degrees of cochlear deafferentation. In the future, these ABR wave I amplitude normative ranges can be used by clinicians to diagnose significant cochlear deafferentation in patients with normal audiograms.

## Acknowledgements

This work was supported by the Department of Veterans Affairs, Veterans Health Administration, Rehabilitation Research and Development Service – Award #C3804-R/I01 RX003804 (to N.F.B) and by resources and facilities at the VA National Center for Rehabilitative Auditory Research (NCRAR) [Center Award #C2361C/I50 RX002361] at the VA Portland Health Care System in Portland, OR. The opinions and assertions presented are private views of the authors and are not to be construed as official or as necessarily reflecting the views of the Department of Veterans Affairs.

## Data Availability Statement

The datafiles generated during and/or analyzed during the current study are available from the corresponding author on reasonable request.

## References

1. Bharadwaj, H. M., Mai, A. R., Simpson, J. M., Choi, I., Heinz, M. G., & Shinn-Cunningham, B. G. (2019). Non-Invasive Assays of Cochlear Synaptopathy – Candidates and Considerations. Neuroscience, 407, 53–66. 10.1016/j.neuroscience.2019.02.031

2. Billings, C. J., Dillard, L. K., Hoskins, Z. B., Penman, T. M., & Reavis, K. M. (2018). A Large-Scale Examination of Veterans with Normal Pure-Tone Hearing Thresholds within the Department of Veterans Affairs. Journal of the American Academy of Audiology, 29(10), 928–935. 10.3766/jaaa.17091

3. Bramhall, N. F. (2021). Use of the auditory brainstem response for assessment of cochlear synaptopathy in humans. The Journal of the Acoustical Society of America, 150(6), 4440. 10.1121/10.0007484

4. Bramhall, N. F., Buran, B. N., & McMillan, G. P. (2025). Associations between physiological indicators of cochlear deafferentation and listening effort in military Veterans with normal audiograms. Hearing Research, 461, 109263. 10.1016/j.heares.2025.109263

5. Bramhall, N. F., Konrad-Martin, D., & McMillan, G. P. (2018). Tinnitus and Auditory Perception After a History of Noise Exposure: Relationship to Auditory Brainstem Response Measures. Ear and Hearing, 39(5), 881–894. 10.1097/AUD.0000000000000544

6. Bramhall, N. F., Konrad-Martin, D., McMillan, G. P., & Griest, S. E. (2017). Auditory Brainstem Response Altered in Humans With Noise Exposure Despite Normal Outer Hair Cell Function. Ear and Hearing, 38(1), e1–e12. 10.1097/AUD.0000000000000370

7. Bramhall, N. F., & McMillan, G. P. (2024). Perceptual Consequences of Cochlear Deafferentation in Humans. Trends in Hearing, 28, 23312165241239541. 10.1177/23312165241239541

8. Bramhall, N. F., McMillan, G. P., Gallun, F. J., & Konrad-Martin, D. (2019). Auditory brainstem response demonstrates that reduced peripheral auditory input is associated with self-report of tinnitus. The Journal of the Acoustical Society of America, 146(5), 3849. 10.1121/1.5132708

9. Bramhall, N. F., McMillan, G. P., & Kampel, S. D. (2021). Envelope following response measurements in young veterans are consistent with noise-induced cochlear synaptopathy. Hearing Research, 408, 108310. 10.1016/j.heares.2021.108310

10. Bramhall, N. F., McMillan, G. P., Kampel, S. D., Heassler, A. E., Whittle, N. K., & Szabo, H. A. (2025). Normative Ranges for Wideband Middle Ear Muscle Reflex Magnitude: Limited Potential for Diagnosing Cochlear Deafferentation. medRxiv, 2025.2009.2012.25335590. 10.1101/2025.09.12.25335590

11. Bramhall, N. F., Theodoroff, S. M., McMillan, G. P., Kampel, S. D., & Buran, B. N. (2023). Associations Between Physiological Correlates of Cochlear Synaptopathy and Tinnitus in a Veteran Population. Journal of Speech, Language, and Hearing Research, 66(11), 4635–4652. 10.1044/2023_JSLHR-23-00234

12. Buran, B. (2015). Auditory-wave-analysis: v1.1. Retrieved January 23, 2025 from http://zenodo.org/record/17365#.VoMCrjbUi70

13. Buran, B. N., Elkins, S., He, W., & Bramhall, N. F. (2025). Predicting cochlear synaptopathy in mice with varying degrees of outer hair cell dysfunction using auditory evoked potentials. bioRxiv, 2025.2005.2009.653157. 10.1101/2025.05.09.653157

14. Carcagno, S., & Plack, C. J. (2020). Effects of age on electrophysiological measures of cochlear synaptopathy in humans. Hearing Research, 396, 108068. 10.1016/j.heares.2020.108068

15. Don, M., & Eggermont, J. J. (1978). Analysis of the click-evoked brainstem potentials in man using high-pass noise masking. The Journal of the Acoustical Society of America, 63(4), 1084–1092.

16. Fletcher, H., & Wegel, R. L. (1922). The Frequency-sensitivity of Normal Ears. Proceedings of the National Academy of Sciences, 8(1), 5–6 2. 10.1073/pnas.8.1.5

17. Gatlin, A. E., & Dhar, S. (2021). History and Lingering Impact of the Arbitrary 25-dB Cutoff for Normal Hearing. American Journal of Audiology, 30(1), 231–234. 10.1044/2020_AJA-20-00181

18. Grant, K. J., Mepani, A. M., Wu, P., Hancock, K. E., de Gruttola, V., Liberman, M. C., & Maison, S. F. (2020). Electrophysiological markers of cochlear function correlate with hearing-in-noise performance among audiometrically normal subjects. Journal of Neurophysiology, 124(2), 418–431. 10.1152/jn.00016.2020

19. Griest-Hines, S. E., Bramhall, N. F., Reavis, K. M., Theodoroff, S. M., & Henry, J. A. (2021). Development and Initial Validation of the Lifetime Exposure to Noise and Solvents Questionnaire in U.S. Service Members and Veterans. American Journal of Audiology, 1–15. 10.1044/2021_AJA-20-00145

20. Gu, J. W., Herrmann, B. S., Levine, R. A., & Melcher, J. R. (2012). Brainstem auditory evoked potentials suggest a role for the ventral cochlear nucleus in tinnitus. Journal of the Association for Research in Otolaryngology, 13(6), 819–833. 10.1007/s10162-012-0344-1

21. Hall, J. W. (2007). New Handbook of Auditory Evoked Responses. Boston, MA: Pearson.

22. Hashimoto, I., Ishiyama, Y., Yoshimoto, T., & Nemoto, S. (1981). Brain-stem auditory-evoked potentials recorded directly from human brain-stem and thalamus. Brain, 104(Pt 4), 841–859.

23. Heassler, A. E., McMillan, G. P., Kampel, S. D., Whittle, N. K., Szabo, H. A., Verhulst, S., … Bramhall, N. F. (2025). Use of Envelope Following Response Normative Ranges for Diagnosing Cochlear Deafferentation. medRxiv. 10.1101/2025.10.24.25338742

24. Henry, J. A., Griest, S., Zaugg, T. L., Thielman, E., Kaelin, C., Galvez, G., & Carlson, K. F. (2015). Tinnitus and hearing survey: a screening tool to differentiate bothersome tinnitus from hearing difficulties. American Journal of Audiology, 24(1), 66–77. 10.1044/2014_AJA-14-0042

25. Hickox, A. E., Larsen, E., Heinz, M. G., Shinobu, L., & Whitton, J. P. (2017). Translational issues in cochlear synaptopathy. Hearing Research, 349, 164–171. 10.1016/j.heares.2016.12.010

26. Johannesen, P. T., Buzo, B. C., & Lopez-Poveda, E. A. (2019). Evidence for age-related cochlear synaptopathy in humans unconnected to speech-in-noise intelligibility deficits. Hearing Research, 374, 35–48.

27. Konrad-Martin, D., Dille, M. F., McMillan, G., Griest, S., McDermott, D., Fausti, S. A., & Austin, D. F. (2012). Age-related changes in the auditory brainstem response. Journal of the American Academy of Audiology, 23(1), 18–35; quiz 74-15. 10.3766/jaaa.23.1.3

28. Kujawa, S. G., & Liberman, M. C. (2009). Adding insult to injury: cochlear nerve degeneration after “temporary” noise-induced hearing loss. Journal of Neuroscience, 29(45), 14077–14085. 10.1523/JNEUROSCI.2845-09.2009

29. Kujawa, S. G., & Liberman, M. C. (2015). Synaptopathy in the noise-exposed and aging cochlea: Primary neural degeneration in acquired sensorineural hearing loss. Hearing Research, 330(Pt B), 191–199. 10.1016/j.heares.2015.02.009

30. Lin, H. W., Furman, A. C., Kujawa, S. G., & Liberman, M. C. (2011). Primary neural degeneration in the Guinea pig cochlea after reversible noise-induced threshold shift. Journal of the Association for Research in Otolaryngology, 12(5), 605–616. 10.1007/s10162-011-0277-0

31. Meikle, M. B., Henry, J. A., Griest, S. E., Stewart, B. J., Abrams, H. B., McArdle, R., … Vernon, J. A. (2012). The tinnitus functional index: development of a new clinical measure for chronic, intrusive tinnitus. Ear and Hearing, 33(2), 153–176. 10.1097/AUD.0b013e31822f67c0

32. Mitchell, C., Phillips, D. S., & Trune, D. R. (1989). Variables affecting the auditory brainstem response: audiogram, age, gender and head size. Hearing Research, 40(1-2), 75–85.

33. Neely, S., & Liu, Z. (1993). EMAV: Otoacoustic emission averager. In Tech Memo No. 17: Boys Town National Research Hospital Omaha.

34. Noble, W., Jensen, N. S., Naylor, G., Bhullar, N., & Akeroyd, M. A. (2013). A short form of the Speech, Spatial and Qualities of Hearing scale suitable for clinical use: the SSQ12. International Journal of Audiology, 52(6), 409–412. 10.3109/14992027.2013.781278

35. Parthasarathy, A., & Kujawa, S. G. (2018). Synaptopathy in the Aging Cochlea: Characterizing Early-Neural Deficits in Auditory Temporal Envelope Processing. Journal of Neuroscience, 38(32), 7108–7119. 10.1523/JNEUROSCI.3240-17.2018

36. Prendergast, G., Guest, H., Munro, K. J., Kluk, K., Leger, A., Hall, D. A., … Plack, C. J. (2017). Effects of noise exposure on young adults with normal audiograms I: Electrophysiology. Hear Res, 344, 68–81. 10.1016/j.heares.2016.10.028

37. Schaette, R., & McAlpine, D. (2011). Tinnitus with a normal audiogram: physiological evidence for hidden hearing loss and computational model. Journal of Neuroscience, 31(38), 13452–13457. 10.1523/JNEUROSCI.2156-11.2011

38. Schmiedt, R. A., Mills, J. H., & Boettcher, F. A. (1996). Age-related loss of activity of auditory-nerve fibers. Journal of Neurophysiology, 76(4), 2799–2803. 10.1152/jn.1996.76.4.2799

39. Sergeyenko, Y., Lall, K., Liberman, M. C., & Kujawa, S. G. (2013). Age-related cochlear synaptopathy: an early-onset contributor to auditory functional decline. Journal of Neuroscience, 33(34), 13686–13694. 10.1523/JNEUROSCI.1783-13.2013

40. Shaheen, L. A., Valero, M. D., & Liberman, M. C. (2015). Towards a Diagnosis of Cochlear Neuropathy with Envelope Following Responses. Journal of the Association for Research in Otolaryngology, 16(6), 727–745. 10.1007/s10162-015-0539-3

41. Shuster, B., Casserly, R., Lipford, E., Olszewski, R., Milon, B., Viechweg, S., … Hertzano, R. (2021). Estradiol Protects against Noise-Induced Hearing Loss and Modulates Auditory Physiology in Female Mice. International Journal of Molecular Sciences, 22(22). 10.3390/ijms222212208

42. Skoe, E., & Tufts, J. (2018). Evidence of noise-induced subclinical hearing loss using auditory brainstem responses and objective measures of noise exposure in humans. Hearing Research, 361, 80–91.

43. Tremblay, K. L., Pinto, A., Fischer, M. E., Klein, B. E., Klein, R., Levy, S., … Cruickshanks, K. J. (2015). Self-Reported Hearing Difficulties Among Adults With Normal Audiograms: The Beaver Dam Offspring Study. Ear and Hearing, 36(6), e290–299. 10.1097/AUD.0000000000000195

44. Valderrama, J. T., Beach, E. F., Yeend, I., Sharma, M., Van Dun, B., & Dillon, H. (2018). Effects of lifetime noise exposure on the middle-age human auditory brainstem response, tinnitus and speech-in-noise intelligibility. Hearing Research, 365, 36–48. 10.1016/j.heares.2018.06.003

45. Valero, M. D., Burton, J. A., Hauser, S. N., Hackett, T. A., Ramachandran, R., & Liberman, M. C. (2017). Noise-induced cochlear synaptopathy in rhesus monkeys (Macaca mulatta). Hearing Research, 353, 213–223. 10.1016/j.heares.2017.07.003

46. Valero, M. D., Hancock, K. E., Maison, S. F., & Liberman, M. C. (2018). Effects of cochlear synaptopathy on middle-ear muscle reflexes in unanesthetized mice. Hearing Research, 363, 109–118. 10.1016/j.heares.2018.03.012

47. Verhulst, S., Altoe, A., & Vasilkov, V. (2018). Computational modeling of the human auditory periphery: Auditory-nerve responses, evoked potentials and hearing loss. Hearing Research, 360, 55–75. 10.1016/j.heares.2017.12.018

48. Wilson, R. H., & McArdle, R. (2013). Characteristics of the audiometric 4,000 Hz notch (744,553 veterans) and the 3,000, 4,000, and 6,000 Hz notches (539,932 veterans). Journal of Rehabilitation Research & Development, 50, 111–132.

49. Wright, E. M., & Royston, P. (1999). Calculating reference intervals for laboratory measurements. Statistical Methods in Medical Research, 8(2), 93–112. 10.1177/096228029900800202

50. Wu, P. Z., Liberman, L. D., Bennett, K., de Gruttola, V., O’Malley, J. T., & Liberman, M. C. (2019). Primary Neural Degeneration in the Human Cochlea: Evidence for Hidden Hearing Loss in the Aging Ear. Neuroscience, 407, 8–20. 10.1016/j.neuroscience.2018.07.053

51. Wu, P. Z., O’Malley, J. T., de Gruttola, V., & Liberman, M. C. (2021). Primary Neural Degeneration in Noise-Exposed Human Cochleas: Correlations with Outer Hair Cell Loss and Word-Discrimination Scores. Journal of Neuroscience, 41(20), 4439–4447. 10.1523/JNEUROSCI.3238-20.2021

